# Relationships between improvement in physical function, pain interference, and mental health in musculoskeletal patients

**DOI:** 10.1101/2023.02.12.23285824

**Authors:** Wei Zhang, Som P Singh, Amdiel Clement, Ryan P Calfee, Janine D Bijsterbosch, Abby L Cheng

## Abstract

**Importance:** Among patients seeking care for musculoskeletal conditions, there is mixed evidence regarding whether traditional, structure-based care is associated with improvement in patients’ mental health.

**Objective:** To determine whether improvements in physical function and pain interference are associated with meaningful improvements in anxiety and depression symptoms among patients seeking musculoskeletal care.

**Design:** Retrospective cohort study from June 22, 2015 to February 9, 2022.

**Setting:** Orthopedic department of a tertiary care US academic medical center.

**Participants:** Consecutive sample of adult patients who presented to the musculoskeletal clinic 4 to 6 times during the study period and completed Patient-Reported Outcomes Measurement Information System (PROMIS) measures as standard care at each visit.

**Exposure:** PROMIS Physical Function and Pain Interference scores.

**Main Outcomes and Measures:** Linear mixed effects models were used to determine whether: 1) PROMIS Anxiety and 2) PROMIS Depression scores improved as a function of improved PROMIS Physical Function or Pain Interference scores, after controlling for age, gender, race, and PROMIS Depression (for the Anxiety model) and PROMIS Anxiety (for the Depression model). Clinically meaningful improvement was defined as ≥3.0 points for PROMIS Anxiety and ≥3.2 points for PROMIS Depression.

**Results:** Among 11,236 patients (mean [SD] age 57 [16] years), 9,706 (86%) were White, and 7,218 (64%) were women. Improvements in physical function (β=-0.14 [95% CI -0.15– -0.13], p<0.001) and pain interference (β=0.26 [0.25-0.26], p<0.001) were each associated with improved anxiety symptoms. To reach a clinically meaningful improvement in anxiety symptoms, an improvement of ≥21 [20-23] PROMIS points on Physical Function or ≥12 [12-12] points on Pain Interference would be required. Improvements in physical function (β=-0.05 [- 0.06– -0.04], p<0.001) and pain interference (β=0.04 [0.04-0.05], p<0.001) were not associated with meaningfully improved depression symptoms.

**Conclusions and Relevance:** In this cohort study, substantial improvements in physical function and pain interference were required for association with any clinically meaningful improvement in anxiety symptoms and were not associated with any meaningful improvement in depression symptoms. Among patients seeking musculoskeletal care, musculoskeletal clinicians and patients cannot assume that addressing physical health will result in improved symptoms of depression or potentially even sufficiently improved symptoms of anxiety.

**Key Points:** *Question:* Among patients seeking musculoskeletal care, are improvements in physical function and pain interference associated with meaningful changes in symptoms of anxiety and depression?

*Findings:* In this large cohort study, improvement by ≥2.3 population-level standard deviations (SD) on PROMIS Physical Function or ≥1.2 SD on PROMIS Pain Interference were required for any association with meaningful improvement in anxiety symptoms. Improvements in physical function and pain interference were not associated with meaningfully improved depression symptoms.

*Meaning:* Musculoskeletal clinicians and patients cannot assume that exclusively addressing the physical aspect of a musculoskeletal condition will improve symptoms of depression or potentially even anxiety.

## Introduction

Physical and mental health have a complex bidirectional relationship, and there is a high prevalence of comorbid physical limitations, pain interference (i.e., “consequences of pain on relevant aspects of a person’s life…including hindered engagement with social, cognitive, emotional, physical, and recreational activities”), and symptoms of anxiety and depression.^1–7^ Clinicians and patients often focus on the treatment of physical concerns, with the hope that mental health related symptoms will naturally improve as physical health improves.^8,9^ This practice may in part be related to unique barriers to accessing mental health care, such as societal stigma regarding mental illness, a lack of financial accessibility to mental health care, and a global shortage of mental health professionals.^10–12^ Furthermore, the structure of medical training is such that clinicians who subspecialize in treating physical impairments do not routinely receive training in addressing the mental health related contributors to and sequelae of physical- and pain-related impairments.^9,13^

Among patients seeking care for musculoskeletal conditions, there is mixed evidence regarding whether treatment of physical conditions is associated with spontaneous improvement in mental health symptoms.^14–16^ Musculoskeletal clinicians also have discrepant opinions regarding whether addressing patients’ mental health falls within their professional role.^9,13,17^ Some musculoskeletal clinicians are interested in additional resources to better address mental health within the musculoskeletal care setting,^18^ but acquisition of these resources has remained challenging without widespread agreement regarding the need for this investment.^13,17,19^ A better understanding of the relationship between physical health and mental health changes can guide musculoskeletal clinicians in: 1) the importance they place on addressing mental health related symptoms as a component of their patient care, and 2) how they counsel patients regarding expectations of their symptom trajectory as their physical impairment is addressed.

The goal of this study was to determine whether, among patients seeking musculoskeletal care, self-reported improvements in physical function and pain interference are each associated with meaningful improvements in self-reported symptoms of anxiety and depression. We hypothesized that clinically meaningful improvement in physical function and pain interference would each be associated with meaningfully improved symptoms of anxiety and depression.

## Methods

This retrospective cohort study included patients who presented to a tertiary academic medical center between June 22, 2015 and February 9, 2022. All study data were extracted from the electronic medical record. Institutional review board approval was granted with a waiver of informed consent.

### Participants

All study participants were adults ≥ 18 years of age who sought evaluation and management of one or more musculoskeletal conditions at an outpatient clinic of the study institution’s orthopedic department. Prior to each clinic evaluation, as standard clinical care, all patients completed Patient-Reported Outcomes Measurement Information System (PROMIS) Computer Adaptive Test (CAT) Anxiety v1.0, Depression v1.0, Physical Function v1.2, and Pain Interference v1.1 measures. Patient visits were excluded from consideration if they were missing scores for any of these measures. To capture time-varying relationships between these PROMIS variables of interest while also maximizing statistical power, our primary cohort included a consecutive sample of patients who had between four and six eligible clinic visits during the study period.

### Exposure and outcome measures

Our exposures of interest were patients’ level of physical function and pain interference over time, which were quantified using their PROMIS CAT Physical Function v1.2 and PROMIS CAT Pain Interference v1.1 scores from each clinic visit, respectively.^7,20–22^ Our outcomes of interest were patients’ symptoms of anxiety and depression, which were quantified using their PROMIS CAT Anxiety v1.0 and Depression v1.0 scores from each clinic visit, respectively.^23,24^

PROMIS is a set of self-reported measures that was developed by the National Institutes of Health to measure multiple domains of health, irrespective of a person’s underlying medical conditions. Scores for each PROMIS measure are normalized to a representative sample of the general US population, with a mean score of 50 and standard deviation of 10. A higher score represents “more” of the domain being assessed, such that a high score on PROMIS Physical Function is favorable, whereas a low score on PROMIS Pain Interference, Anxiety, and Depression is favorable. Clinically meaningful effect sizes for symptom improvement were defined as at least 3.0 points for PROMIS Anxiety and 3.2 points for PROMIS Depression, which, based on the literature, are the minimum clinically important differences among patients with musculoskeletal pain that also exceed the standard error of measurement for each PROMIS CAT at the study institution.^26,27^

### Confounding variables

Patients’ age, self-reported gender, and self-reported race were also available in the medical record and were included in all statistical models to account for potential confounding effects.

### Generalizability analyses

To assess the generalizability of our findings to patients who may have less chronic and/or less severe musculoskeletal conditions, a second cohort was also analyzed which consisted of a consecutive sample of patients who had only three clinic visits during the six-year study period (as compared to our primary cohort which had four to six visits). These patients were not included in our primary cohort because our goal was to optimally capture time-varying relationships between the measures of interest, so our primary cohort included patients who had more than three visits during the study period.

### Statistical Analysis

Linear mixed effects models were used for all analyses and accounted for both fixed and random effects, which therefore controlled for individual variability in relationships between exposure and outcome measures. Separate models analyzed PROMIS Anxiety and PROMIS Depression. First, we tested for a main effect of PROMIS Physical Function on PROMIS Anxiety and PROMIS Depression, with adjustment for potential confounding effects from age, gender, and race (categorized as White versus non-White because the sample was predominantly White). Because anxiety and depression symptoms are frequently comorbid and highly correlated,^28^ we also adjusted for PROMIS Depression and PROMIS Anxiety scores as covariates in the respective models in order to account for shared variance and to test for effects of physical function specific to either disorder (i.e., specific main effects models). For all models, we included a random intercept for each patient to account for individual-level variance (e.g., irregular time interval between visits) and a random intercept for each clinic visit to account for variability in the number of visits. If a model failed to converge even after maximizing the number of iterations (i.e., data did not support the model specifications), the random intercept for each clinic visit was dropped. All resulting statistical models converged. Calculated β coefficients of less than 0.06 were considered to indicate negligible relationships because they would represent changes in PROMIS Anxiety or Depression scores that would not reach our predefined minimum clinically meaningful thresholds, even if the greatest possible score change in PROMIS Physical Function or Pain Interference was achieved.

Next, to focus on patients who, according to our hypothesis, would be most likely to achieve clinically meaningful improvement in symptoms of anxiety and depression, we repeated all analyses on the subgroup of patients whose physical function meaningfully improved (i.e., five-point score increase in PROMIS Physical Function) between their first and last clinic visit during the study period (Figure S1). While the precise threshold for “meaningful improvement” varies based on the patient population and is not universally agreed upon, five points was chosen as the cutoff value for this study because it corresponds to a moderate effect size change (i.e., 0.5 standard deviations) and has repeatedly been found to represent a clinically important difference across various orthopedic patient populations.^26,29,30^

We then repeated all statistical procedures using PROMIS Pain Interference as the exposure measure (i.e., predictor), rather than PROMIS Physical Function. Meaningfully improved pain interference was defined as a five-point decrease in PROMIS Pain Interference.^26,31,32^ Of note, shared variance between PROMIS Physical Function and Pain Interference was not accounted for in any of the models because identifying the unique contribution from each of these domains was not the purpose of this study.

All statistical procedures were also repeated in follow-up tests using the generalizability cohort of patients who only had three clinic visits during the study period (as compared to our primary cohort which had four to six visits). These tests were conducted to examine whether findings from our primary cohort were biased due to specific characteristics of the cohort (e.g., potentially more chronic musculoskeletal conditions). The sample size of each cohort was determined by the availability of eligible patients. Given the large available sample size, the relatively small proportion of patients with missing PROMIS scores were excluded from analysis based on the study eligibility criteria. P-values were derived from F-tests using Satterthwaite’s methods.^33^ To account for multiple comparisons, false discovery rate (FDR) corrections were applied to all the models. All statistical analyses were conducted using R v4.10 (Vienna, Austria). Linear mixed effects models were conducted using the lmerTest package,^34^ and FDR corrections were applied using the “p.adjust” function and implementing Benjamini and Hochberg’s approach.^35^

## Results

### Demographics

Of 87,490 patients who were evaluated at the study institution during the study period, 11,236 were eligible for inclusion in the primary cohort (51,569 total visits) (Figure 1). This cohort had a mean [SD] age 58 [16] years, 9,706 (86%) were White, and 7,218 (64%) were women (Table 1). Demographic characteristics were similar among the subgroup of patients who achieved meaningfully improved physical function and/or pain interference during the study period.

**Figure 1:**
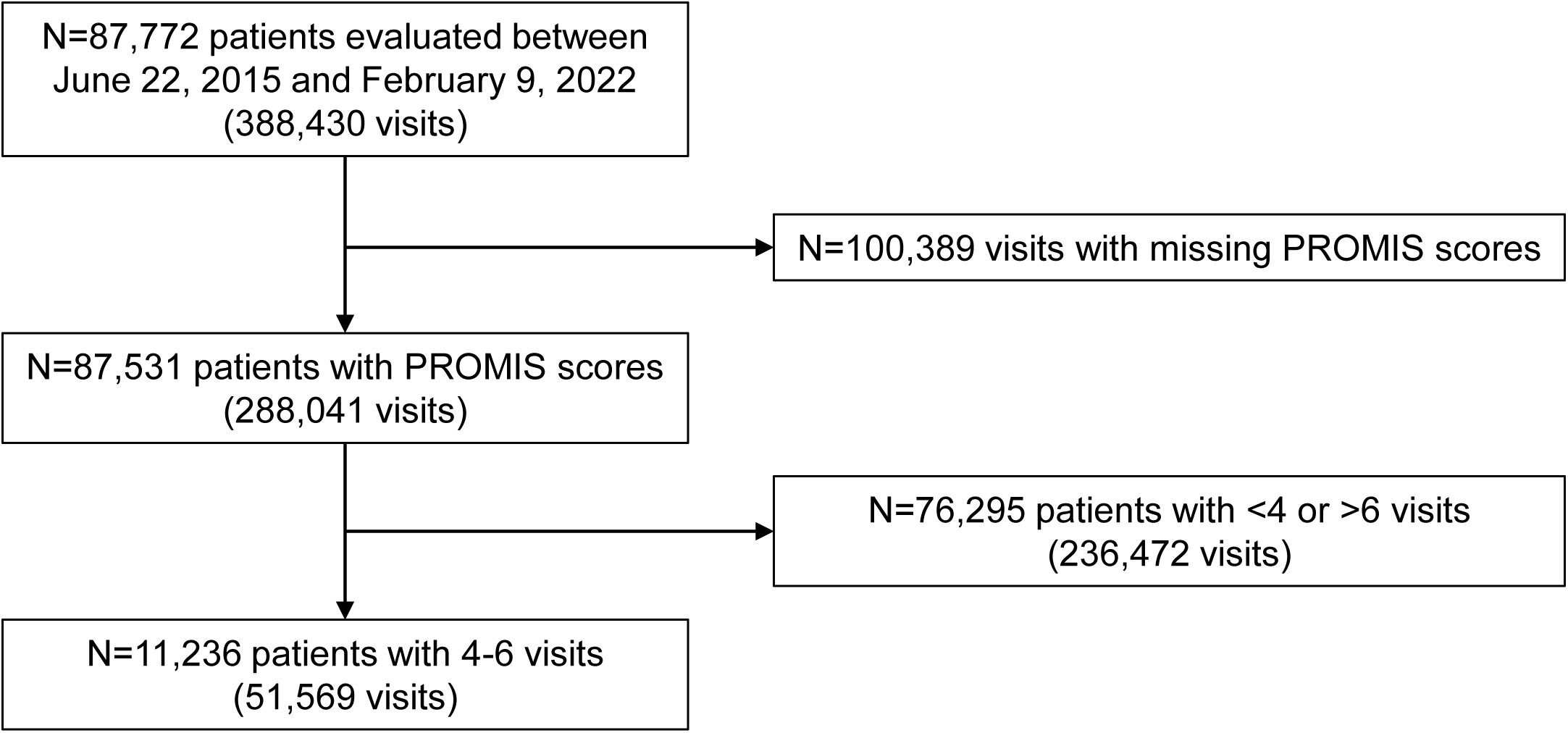
Inclusion flowsheet. Abbreviation: PROMIS (Patient-Reported Outcomes Measurement Information System.

**Table 1:** Patient characteristics.

|  | **Primary cohort**<br>(N=11,236) | **Physical Function**<br>**Improved<sup>a</sup>**<br>(N=1,672) | **Pain Interference**<br>**Improved<sup>a</sup>**<br>(N=1,391) |
| --- | --- | --- | --- |
| Age, Mean (SD) | 58 (16) | 57 (16) | 58 (15) |
| Women, N (%) | 7,218 (64) | 1,067 (64) | 928 (67) |
| Race, N (%) |  |  |  |
| White | 9,706 (86) | 1,455 (87) | 1,203 (87) |
| Black or African American | 1,288 (12) | 190 (11) | 161 (12) |
| Asian | 120 (1) | 15 (< 1) | 15 (1) |
| American Indian or Alaska Native | 26 (< 1) | 1 (< 1) | 2 (< 1) |
| Other Pacific Islander | 15 (< 1) | 2 (< 1) | 1 (< 1) |
| Multi-racial | 39 (< 1) | 5 (< 1) | 1 (< 1) |
| Unable to answer | 1 (< 1) | 0 (0) | 1 (< 1) |
| Declined | 41 (< 1) | 4 (< 1) | 7 (< 1) |

<sup>a</sup>“Improved” was defined as a five-point favorable change in patients’ Patient-Reported
Outcomes Measurement Information System (PROMIS) scores from the first to last clinic visit
during the study period (i.e., five-point score increase in PROMIS Physical Function, five-point
score decrease in PROMIS Pain Interference).

### Effects on anxiety

For the primary cohort after adjusting for age, gender, race, and depression symptoms, improvements in physical function (β=-0.14 [95% CI -0.15 to -0.13], p_fdr_<0.001) and pain interference (β=0.26 [0.25 to 0.26], p_fdr_<0.001) were each associated with statistically and meaningfully improved anxiety symptoms (Figure 2, Table 2). To reach a clinically meaningful improvement in anxiety symptoms of at least 3.0 PROMIS Anxiety points, an improvement of ≥21 [20 to 23] PROMIS points on Physical Function or ≥12 [12 to 12] points on Pain Interference would be required (calculated as 3.0/β).

**Figure 2:**
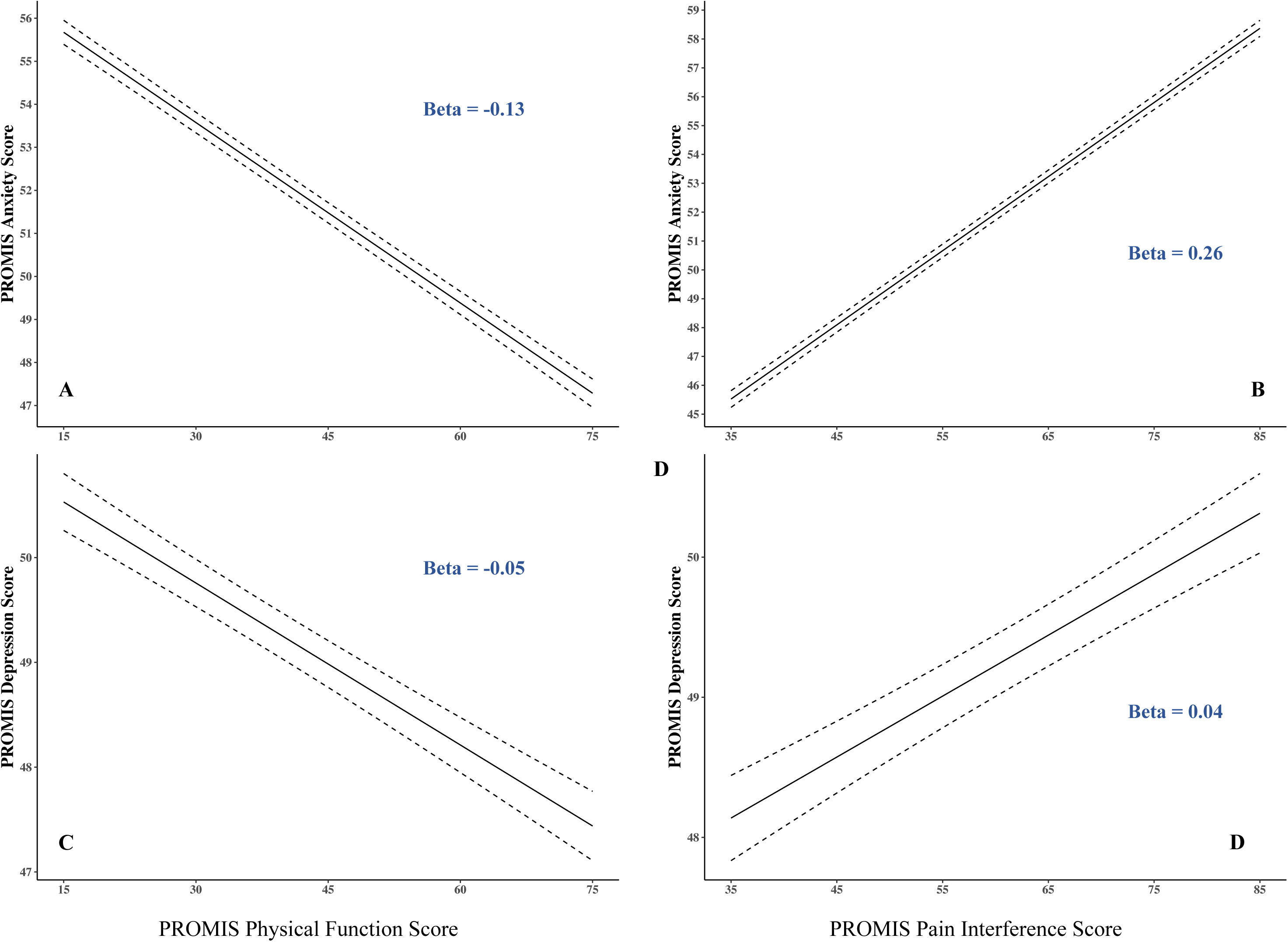
Predicted PROMIS mental health scores as a function of PROMIS physical health scores. Predicted PROMIS Anxiety (A and B) and Depression (C and D) scores as a function of PROMIS Physical Function (A and C) and Pain Interference (B and D) scores across all treatment visits, after adjusting for age, gender, race, and shared variance between PROMIS Depression and Anxiety. Dashed lines indicate 95% confidence intervals. Note the different Y-axis scales in each panel.

**Table 2:** Models testing for an effect of PROMIS Physical Function and Pain Interference on PROMIS Anxiety.

| Predictor | PROMIS Physical Function |  |  | PROMIS Pain Interference |  |  |
| --- | --- | --- | --- | --- | --- | --- |
| | Estimate ($\beta$) | 95% CI | *p* | Estimate ($\beta$) | 95% CI | *p* |
| (Intercept) | 29.83 | 29.24 to 30.42 | <0.001 | 10.45 | 9.89 to 11.01 | <0.001 |
| PROMIS Physical Function | -0.14 | -0.15 to -0.13 | <0.001 | NI | NI | NI |
| PROMIS Pain Interference | NI | NI | NI | 0.26 | 0.25 to 0.26 | <0.001 |
| PROMIS Depression | 0.58 | 0.58 to 0.59 | <0.001 | 0.54 | 0.54 to 0.55 | <0.001 |
| Age (per year) | -0.02 | -0.02 to -0.01 | <0.001 | -0.01 | -0.02 to -0.01 | <0.001 |
| Gender (Man) | -0.03 | -0.20 to 0.13 | 0.70 | -0.12 | -0.28 to 0.03 | 0.13 |
| Race (White) | -2.23 | -2.46 to -2.00 | <0.001 | -1.79 | -2.01 to -1.57 | <0.001 |
| Visit 2 | 2.44 | 2.32 to 2.57 | <0.001 | 2.29 | 2.16 to 2.41 | <0.001 |
| Visit 3 | 3.92 | 3.79 to 4.05 | <0.001 | 3.65 | 3.53 to 3.78 | <0.001 |
| Visit 4 | 5.68 | 5.55 to 5.81 | <0.001 | 5.28 | 5.15 to 5.41 | <0.001 |
| Visit 5 | 6.50 | 6.33 to 6.67 | <0.001 | 6.05 | 5.88 to 6.22 | <0.001 |
| Visit 6 | 7.59 | 7.32 to 7.85 | <0.001 | 7.01 | 6.75 to 7.27 | <0.001 |
| **Random Effects** |  |  |  |  |  |  |
| $\sigma^2$ | 22.95 | - | - | 21.46 | - | - |
| $\tau_{00,id}$ | 12.76 | - | - | 12.15 | - | - |
| ICC | 0.36 | - | - | 0.36 | - | - |
| $N_{id}$ | 11,236 | - | - | 11,236 | - | - |
| Observations | 51,569 | - | - | 51,569 | - | - |
| Marginal $R^2$ / Conditional $R^2$ | 0.561 / 0.718 | - | - | 0.585 / 0.735 | - | - |

Abbreviations: PROMIS (Patient-Reported Outcomes Measurement Information System), NI (Not included in the model), $\sigma^2$ (random effect variance), $\tau_{00,id}$ (random intercept for each individual patient), ICC (Intra-Class-Correlation coefficient), $N_{id}$ (number of patients), Observations (number of datapoints for all included patients across all visits).

Among the subgroup of patients who achieved meaningfully improved physical function or pain interference during the study period (of at least five PROMIS points), these relationships were similar (physical function β=-0.11 [-0.13 to -0.09], p_fdr_<0.001; pain interference β=0.22 [0.19 to 0.24], p_fdr_<0.001) (Table S1). For this subgroup to reach a clinically meaningful improvement in anxiety symptoms, a somewhat greater improvement of ≥27 [23 to 33] PROMIS points on Physical Function or ≥14 [13 to 16] points on Pain Interference would be required.

### Effects on depression

For the primary cohort after adjusting for age, gender, race, and anxiety symptoms, improvements in physical function (β=-0.05 [-0.06 to -0.04], p_fdr_<0.001) and pain interference (β=0.04 [0.04 to 0.05], p_fdr_<0.001) were each associated with statistically but not meaningfully improved depression symptoms (Figure 2, Table 3). That is, to reach a clinically meaningful improvement in depression symptoms of at least 3.2 PROMIS Depression points, an improvement of ≥64 [53 to 80] PROMIS points on Physical Function or ≥64 [64 to 80] points on Pain Interference would be required (calculated as 3.2/β), which is not possible based on the actual score ranges of these PROMIS measures.

**Table 3:** Models testing for an effect of PROMIS Physical Function and Pain Interference on PROMIS Depression.

| Predictor | PROMIS Physical Function |  |  | PROMIS Pain Interference |  |  |
| --- | --- | --- | --- | --- | --- | --- |
| | Estimate ($\beta$) | 95% CI | *p* | Estimate ($\beta$) | 95% CI | *p* |
| (Intercept) | 17.34 | 16.71 to 17.96 | <0.001 | 12.65 | 12.08 to 13.22 | <0.001 |
| PROMIS Physical Function | -0.05 | -0.06 to -0.04 | <0.001 | NI | NI | NI |
| PROMIS Pain Interference | NI | NI | NI | 0.04 | 0.04 to 0.05 | <0.001 |
| PROMIS Anxiety | 0.64 | 0.64 to 0.65 | <0.001 | 0.64 | 0.63 to 0.65 | <0.001 |
| Age (per year) | -0.01 | -0.02 to -0.01 | <0.001 | -0.01 | -0.01 to -0.01 | <0.001 |
| Gender (Man) | -0.17 | -0.32 to -0.01 | 0.032 | -0.21 | -0.36 to -0.06 | 0.006 |
| Race (White) | 0.61 | 0.30 to 0.73 | <0.001 | 0.66 | 0.48 to 0.88 | <0.001 |
| Visit 2 | -0.20 | -0.33 to -0.06 | 0.005 | -0.18 | -0.32 to -0.04 | 0.01 |
| Visit 3 | -0.29 | -0.43 to -0.15 | <0.001 | -0.27 | -0.41 to -0.13 | <0.001 |
| Visit 4 | -0.46 | -0.61 to -0.31 | <0.001 | -0.44 | -0.59 to -0.29 | <0.001 |
| Visit 5 | -0.45 | -0.64 to -0.26 | <0.001 | -0.42 | -0.61 to -0.23 | <0.001 |
| Visit 6 | -0.26 | -0.55 to -0.03 | 0.079 | -0.24 | -0.53 to -0.06 | 0.11 |
| **Random Effects** |  |  |  |  |  |  |
| $\sigma^2$ | 26.39 | - | - | 26.40 | - | - |
| $\tau_{00,id}$ | 9.38 | - | - | 9.51 | - | - |
| ICC | 0.26 | - | - | 0.26 | - | - |
| $N_{id}$ | 11,236 | - | - | 11,236 | - | - |
| Observations | 51,569 | - | - | 51,569 | - | - |
| Marginal $R^2$ / Conditional $R^2$ | 0.514 / 0.641 | - | - | 0.511 / 0.641 | - | - |

Abbreviations: PROMIS (Patient-Reported Outcomes Measurement Information System), NI (Not included in the model), $\sigma^2$ (random effect variance), $\tau_{00,id}$ (random intercept for each individual patient), ICC (Intra-Class-Correlation coefficient), $N_{id}$ (number of patients), Observations (number of datapoints for all included patients across all visits).

Among the subgroup of patients who achieved meaningfully improved physical function or pain interference during the study period (of at least five PROMIS points), these relationships were unchanged (physical function β=-0.03 [95% CI -0.05 to -0.02], p_fdr_<0.001; pain interference β=0.04 [0.02 to 0.06], p_fdr_<0.001) (Table S2). Specifically, the models for this subgroup still suggest that it would not be possible to reach a clinically meaningful improvement in depression symptoms of 3.2 PROMIS points due only to improvements in physical function or pain interference.

### Generalizability analysis

Compared to our primary cohort of patients who had at least four clinic visits during the study period, no meaningful differences in baseline characteristics (Table S3) or physical/mental health relationships (Table S4 and Table S5) were observed in our generalizability cohort of patients who only had three visits during the six-year study period.

## Discussion

Our findings suggest that on a departmental level, improvements in physical function and pain interference independently contribute to improvements in anxiety symptoms. Nevertheless, to be associated with clinically meaningful improvements in anxiety symptoms, substantial changes of at least 2.1 physical function standard deviations (of the U.S. population) or at least 1.2 pain interference standard deviations are required. Furthermore, even drastic improvements in physical function or pain interference are not independently associated with meaningful improvements in depression symptoms. These study findings were consistent, even when considering 1) all patients versus only patients who achieved meaningfully improved physical function or pain interference during the study period, and 2) patients with 4-6 clinic visits versus only 3 visits during the study period.

There is consistent evidence for the negative impact of *preexisting* anxiety and depression symptoms on physical health related outcomes after musculoskeletal treatment.^36–39^ In contrast, this study adds to the emerging literature that elucidates the association between improved physical health and subsequent symptoms of anxiety and depression.^14–16^ So far, this emerging literature has been mixed. For example, use of mental health related medications and psychotherapy was found to decrease in a cohort of young adults after undergoing hip surgery,^15^ whereas symptoms of depression did not meaningfully change after a different cohort of patients underwent surgical or non-surgical treatment for a variety of orthopedic hand conditions.^16^ It is possible that some musculoskeletal treatments may improve symptoms of anxiety and depression in the short-term for patients who, for instance, may have situational/state (rather than trait) anxiety or depression.^40^ This could especially be true for elective orthopedic procedures which have a high success rate and are typically only performed on people who are generally healthy at baseline.

However, in contrast to these previous studies, this study was not designed to assess the effectiveness of a *specific* musculoskeletal treatment for a *specific* patient population. Rather, this study was designed to identify broad associations between physical and mental health trajectories over a long time period, regardless of a patient’s precise musculoskeletal condition(s) or structure-based treatment (e.g., surgery, injection, physical therapy, etc). Our findings suggest that over the long-term, musculoskeletal clinicians should be aware that improvements in physical function and/or pain interference are not necessarily associated with meaningful and sustained improvements in symptoms of anxiety or especially depression. Furthermore, because preexisting symptoms of anxiety and depression are associated with worse orthopedic outcomes, we advocate for musculoskeletal clinicians to be equipped with the training and referral resources to address mental health as part of patient counseling and the musculoskeletal treatment plan.^13^

### Limitations

Although this study had key strengths including the large sample size and relatively long follow-up duration of over six years, there were also several limitations. First, this was an observational study, which limits the causality we can attribute to the relationships between physical and mental health that we identified. Second, the patient cohort had limited racial diversity. Third, additional sociodemographic and clinical variables were not available to be included as possible confounders (e.g., financial considerations, social support, and medical comorbidities). Based on our generalizability analysis and previous work,^14^ it is unlikely that our findings are specific to patients with more severe, more chronic, more refractory, and/or multiple musculoskeletal conditions (i.e., patients who had at least four clinic visits during the study period). However, it is possible that other patient, diagnosis, and treatment characteristics could influence the relationships we identified (e.g. traumatic versus degenerative conditions, spine versus peripheral joint conditions, definitive versus palliative treatment intent, etc).

## Conclusion

This large cohort study suggests that over the course of several years, improvements in physical function and pain interference may be associated with improvements in symptoms of anxiety but not of depression. Furthermore, substantial improvements in physical function and pain interference are required in order to reach clinically meaningful associations with improvement in anxiety symptoms. Therefore, musculoskeletal clinicians and patients cannot assume that exclusively structure-based treatment of a musculoskeletal condition will necessarily result in improved symptoms of depression or potentially even anxiety. We advocate for clinicians to thoughtfully and intentionally address the mental health related contributors to, and sequelae of, musculoskeletal conditions when counseling patients and creating person-centered treatment plans. Further investigation is needed to identify methods of addressing mental health in the context of musculoskeletal care that are both feasible and effective.

## Data Availability

All data produced in the present study are available upon reasonable request to the authors.

## Author Contributions

Dr. Cheng had full access to all the data in the study and takes responsibility for the integrity of the data and the accuracy of the data analysis.

*Concept and design:* Cheng, Calfee, Zhang.

*Acquisition and analysis:* Cheng, Zhang.

*Interpretation of data:* All authors.

*Drafting of the manuscript:* Zhang, Singh, Clements, Cheng.

*Critical revision of the manuscript for important intellectual content:* All authors.

*Obtained funding:* Cheng.

*Administrative, technical, or material support:* Cheng.

*Supervision:* Cheng.

## Previous Presentation of Information

The findings described in this manuscript have not previously been presented.

## Additional Contributions

(Not applicable.)

---

**Table S1:** Effect of PROMIS Physical Function and Pain Interference on PROMIS Anxiety, among patients with clinically improved physical function or pain interference.

| Predictor | PROMIS Physical Function |  |  | PROMIS Pain Interference |  |  |
| --- | --- | --- | --- | --- | --- | --- |
| | Estimate ($\beta$) | 95% CI | *p* | Estimate ($\beta$) | 95% CI | *p* |
| (Intercept) | 25.82 | 24.32 to 27.32 | <0.001 | 9.11 | 6.04 to 12.18 | <0.001 |
| PROMIS Physical Function | -0.11 | -0.13 to -0.09 | <0.001 | NI | NI | NI |
| PROMIS Pain Interference | NI | NI | NI | 0.22 | 0.19 to 0.24 | <0.001 |
| PROMIS Depression | 0.65 | 0.63 to 0.66 | <0.001 | 0.63 | 0.61 to 0.65 | <0.001 |
| Age (per year) | -0.02 | -0.03 to -0.01 | <0.005 | -0.02 | -0.03 to 0.001 | 0.008 |
| Gender = Man | 0.17 | -0.26 to 0.61 | 0.432 | -0.20 | -0.66 to 0.26 | 0.39 |
| Race = White | -2.38 | -3.00 to -1.76 | <0.001 | -1.50 | -2.13 to -0.86 | <0.001 |
| Visit 2 | 2.68 | 2.32 to 3.04 | <0.001 | 2.30 | -1.25 to 5.84 | 0.20 |
| Visit 3 | 3.58 | 3.22 to 3.94 | <0.001 | 2.91 | -0.63 to 6.46 | 0.11 |
| Visit 4 | 5.14 | 4.76 to 5.52 | <0.001 | 4.60 | 1.05 to 8.14 | 0.011 |
| Visit 5 | 6.12 | 5.62 to 6.62 | <0.001 | 5.59 | 2.02 to 9.15 | 0.002 |
| Visit 6 | 7.34 | 6.57 to 8.12 | <0.001 | 6.44 | 2.82 to 10.06 | <0.001 |
| **Random Effects** |  |  |  |  |  |  |
| $\sigma^2$ | 25.80 | - | - | 25.62 | - | - |
| $\tau_{00,id}$ | 12.93 | - | - | 11.06 | - | - |
| $\tau_{00,visit}$ | NI | - | - | 1.61 | - | - |
| ICC | 0.33 | - | - | 0.33 | - | - |
| $N_{id}$ | 1,672 | - | - | 1,391 | - | - |
| $N_{visit}$ | NI | - | - | 6 | - | - |
| Observations | 7,652 | - | - | 6,366 | - | - |
| Marginal $R^2$ / Conditional $R^2$ | 0.532 / 0.688 | - | - | 0.524 / 0.681 | - | - |

“Clinically improved” physical function or pain interference was defined as a five-point favorable change in patients’ Patient-Reported Outcomes Measurement Information System (PROMIS) scores from the first to last clinic visit during the study period (i.e., five-point score increase in PROMIS Physical Function, five-point score decrease in PROMIS Pain Interference).
Abbreviations: PROMIS (Patient-Reported Outcomes Measurement Information System), NI (Not included in the model), $\sigma^2$ (random effect variance), $\tau_{00,id}$ (random intercept for each individual patient), $\tau_{00,visit}$ (random intercept for each clinic visit), ICC (Intra-Class-Correlation coefficient), $N_{id}$ (number of patients), $N_{visit}$ (number of clinic visits), Observations (number of datapoints for all included patients across all visits).

**Table S2:** Effect of PROMIS Physical Function and Pain Interference on PROMIS Depression, among patients with clinically improved physical function or pain interference.

| Predictor | PROMIS Physical Function |  |  | PROMIS Pain Interference |  |  |
| --- | --- | --- | --- | --- | --- | --- |
| | Estimate ($\beta$) | 95% CI | *p* | Estimate ($\beta$) | 95% CI | *p* |
| (Intercept) | 16.13 | 14.60 to 17.65 | <0.001 | 13.10 | 9.71 to 16.49 | <0.001 |
| PROMIS Physical Function | -0.03 | -0.05 to -0.02 | <0.001 | NI | NI | NI |
| PROMIS Pain Interference | NI | NI | NI | 0.04 | 0.02 to 0.06 | <0.001 |
| PROMIS Anxiety | 0.65 | 0.63 to 0.67 | <0.001 | 0.64 | 0.62 to 0.66 | <0.001 |
| Age (per year) | 0.00 | -0.01 to 0.01 | 0.70 | -0.01 | -0.02 to 0.01 | 0.34 |
| Gender (Man) | -0.56 | -0.97 to -0.16 | 0.005 | -0.04 | -0.49 to 0.42 | 0.80 |
| Race (White) | 0.26 | -0.32 to 0.84 | 0.53 | 0.29 | -0.35 to 0.93 | 0.51 |
| Visit 2 | -0.18 | -0.55 to 0.18 | 0.33 | -0.09 | -4.18 to 4.01 | 0.97 |
| Visit 3 | -0.07 | -0.45 to 0.30 | 0.70 | -0.08 | -4.17 to 4.01 | 0.97 |
| Visit 4 | -0.19 | -0.60 to 0.21 | 0.35 | -0.10 | -4.19 to 4.00 | 0.96 |
| Visit 5 | -0.43 | -0.95 to 0.09 | 0.11 | -0.44 | -4.55 to 3.67 | 0.83 |
| Visit 6 | -0.82 | -1.62 to -0.02 | 0.045 | -0.38 | -4.54 to 3.78 | 0.86 |
| **Random Effects** |  |  |  |  |  |  |
| $\sigma^2$ | 26.63 | - | - | 26.14 | - | - |
| $\tau_{00,id}$ | 10.42 | - | - | 10.92 | - | - |
| $\tau_{00,visit}$ | NI | - | - | 2.16 | - | - |
| ICC | 0.28 | - | - | 0.33 | - | - |
| $N_{id}$ | 1,672 | - | - | 1,391 | - | - |
| $N_{visit}$ | NI | - | - | 6 | - | - |
| Observations | 7,625 | - | - | 6,366 | - | - |
| Marginal $R^2$ / Conditional $R^2$ | 0.502 / 0.642 | - | - | 0.468 / 0.646 | - | - |

“Clinically improved” physical function or pain interference was defined as a five-point favorable change in patients’ Patient-Reported Outcomes Measurement Information System (PROMIS) scores from the first to last clinic visit during the study period (i.e., five-point score increase in PROMIS Physical Function, five-point score decrease in PROMIS Pain Interference).
Abbreviations: PROMIS (Patient-Reported Outcomes Measurement Information System), NI (Not included in the model), $\sigma^2$ (random effect variance), $\tau_{00,id}$ (random intercept for each individual patient), $\tau_{00,visit}$ (random intercept for each clinic visit), ICC (Intra-Class-Correlation coefficient), $N_{id}$ (number of patients), $N_{visit}$ (number of clinic visits), Observations (number of datapoints for all included patients across all visits).

**Table S3:** Patient characteristics of the generalizability cohort who had three clinic visits during the study period.

|  | **Generalizability cohort**<br>(N=11,501) |
| --- | --- |
| Age, Mean (SD) | 56 (16) |
| Women, N (%) | 6,966 (61) |
| Race, N (%) |  |
| White | 9,864 (86) |
| Black or African American | 1,342 (12) |
| Asian | 171 (2) |
| American Indian or Alaska Native | 18 (< 1) |
| Other Pacific Islander | 12 (< 1) |
| Multi-racial | 47 (< 1) |
| Unable to answer | 5 (< 1) |
| Declined | 42 (< 1) |

**Table S4:**
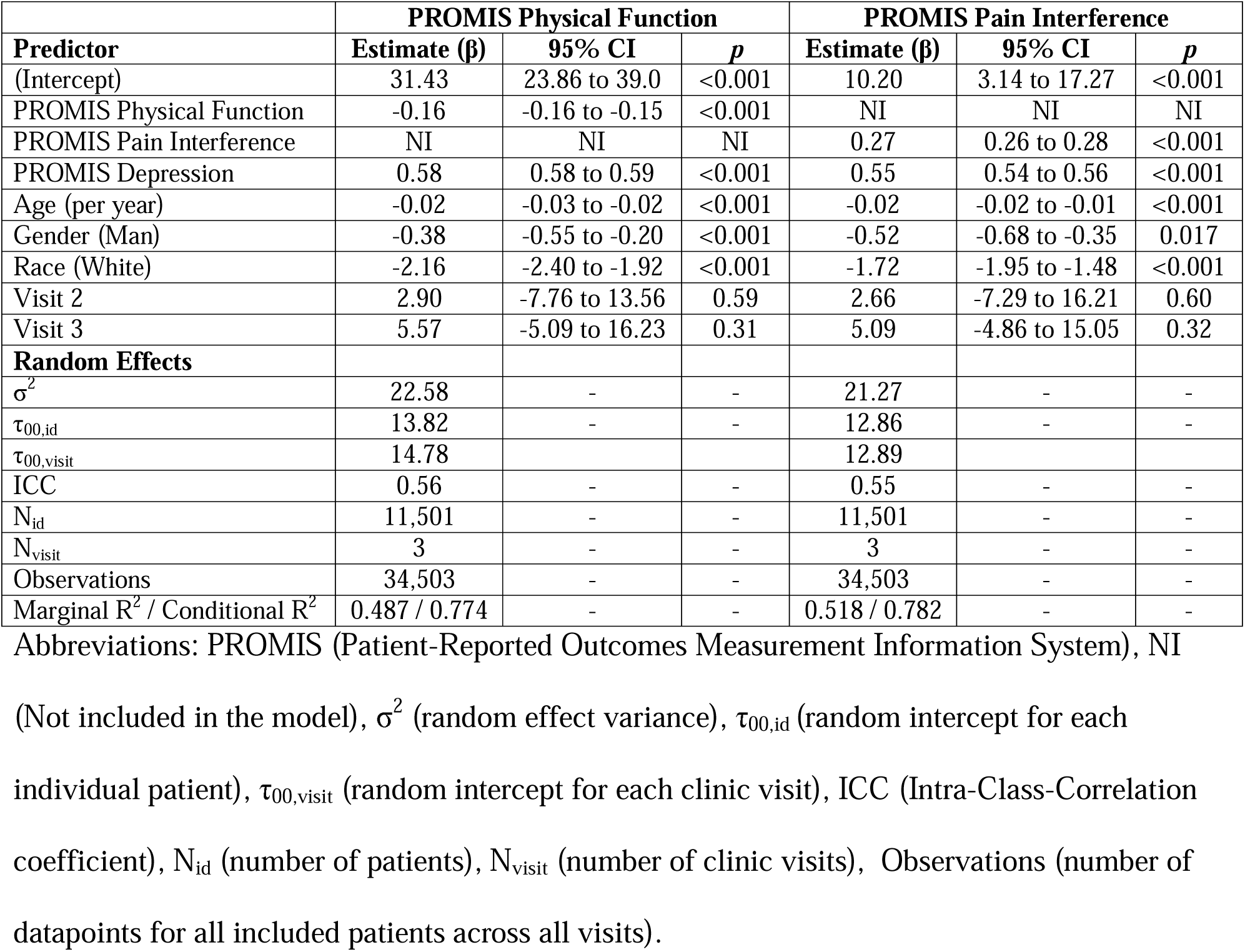
Effect of PROMIS Physical Function and Pain Interference on PROMIS Anxiety in the generalizability cohort. In patients who only had three visits during the study period, after adjusting for age, gender, race, and depression symptoms, improvements in physical function (β=-0.16 [95% CI -0.16 to -0.15], p_fdr_<0.001) and pain interference (β=0.27 [0.26 to 0.28], pŗ_dr_<0.001) were each associated with statistically and meaningfully improved anxiety symptoms (Table S4). To reach a clinically meaningful improvement in anxiety symptoms of at least 3.0 PROMIS Anxiety points, an improvement of ≥19 [19 to 20] PROMIS points on Physical Function or ≥11 [11 to 12] points on Pain Interference would be required (calculated as 3.0/β).

| Predictor | PROMIS Physical Function |  |  | PROMIS Pain Interference |  |  |
| --- | --- | --- | --- | --- | --- | --- |
| | Estimate ($\beta$) | 95% CI | *p* | Estimate ($\beta$) | 95% CI | *p* |
| (Intercept) | 31.43 | 23.86 to 39.0 | <0.001 | 10.20 | 3.14 to 17.27 | <0.001 |
| PROMIS Physical Function | -0.16 | -0.16 to -0.15 | <0.001 | NI | NI | NI |
| PROMIS Pain Interference | NI | NI | NI | 0.27 | 0.26 to 0.28 | <0.001 |
| PROMIS Depression | 0.58 | 0.58 to 0.59 | <0.001 | 0.55 | 0.54 to 0.56 | <0.001 |
| Age (per year) | -0.02 | -0.03 to -0.02 | <0.001 | -0.02 | -0.02 to -0.01 | <0.001 |
| Gender (Man) | -0.38 | -0.55 to -0.20 | <0.001 | -0.52 | -0.68 to -0.35 | 0.017 |
| Race (White) | -2.16 | -2.40 to -1.92 | <0.001 | -1.72 | -1.95 to -1.48 | <0.001 |
| Visit 2 | 2.90 | -7.76 to 13.56 | 0.59 | 2.66 | -7.29 to 16.21 | 0.60 |
| Visit 3 | 5.57 | -5.09 to 16.23 | 0.31 | 5.09 | -4.86 to 15.05 | 0.32 |
| **Random Effects** |  |  |  |  |  |  |
| $\sigma^2$ | 22.58 | - | - | 21.27 | - | - |
| $\tau_{00,id}$ | 13.82 | - | - | 12.86 | - | - |
| $\tau_{00,visit}$ | 14.78 | - | - | 12.89 | - | - |
| ICC | 0.56 | - | - | 0.55 | - | - |
| $N_{id}$ | 11,501 | - | - | 11,501 | - | - |
| $N_{visit}$ | 3 | - | - | 3 | - | - |
| Observations | 34,503 | - | - | 34,503 | - | - |
| Marginal $R^2$ / Conditional $R^2$ | 0.487 / 0.774 | - | - | 0.518 / 0.782 | - | - |

Abbreviations: PROMIS (Patient-Reported Outcomes Measurement Information System), NI (Not included in the model), $\sigma^2$ (random effect variance), $\tau_{00,id}$ (random intercept for each individual patient), $\tau_{00,visit}$ (random intercept for each clinic visit), ICC (Intra-Class-Correlation coefficient), $N_{id}$ (number of patients), $N_{visit}$ (number of clinic visits), Observations (number of datapoints for all included patients across all visits).

**Table S5:** Effect of PROMIS Physical Function and Pain Interference on PROMIS Depression in the generalizability cohort. After adjusting for age, gender, race, and anxiety symptoms, improvements in physical function (β=-0.04 [-0.05 to -0.03], p_fdr_<0.001) and pain interference (β=0.03 [0.02 to 0.04], p_f_dľ<0.001) were associated with statistically but not meaningfully improved depression symptoms (Table S5). That is, to reach a clinically meaningful improvement in depression symptoms of at least 3.2 PROMIS Depression points, an improvement of ≥80 [64 to 107] PROMIS points on Physical Function or ≥107 [80 to 160] points on Pain Interference would be required (calculated as 3.2/β), which is not possible based on the actual score ranges of these PROMIS measures.

| Predictor | PROMIS Physical Function |  |  | PROMIS Pain Interference |  |  |
| --- | --- | --- | --- | --- | --- | --- |
| | Estimate ($\beta$) | 95% CI | *p* | Estimate ($\beta$) | 95% CI | *p* |
| (Intercept) | 14.53 | 12.73 to 16.33 | 0.007 | 11.13 | 9.83 to 12.43 | <0.001 |
| PROMIS Physical Function | -0.04 | -0.05 to -0.03 | <0.001 | NI | NI | NI |
| PROMIS Pain Interference | NI | NI | NI | 0.03 | 0.02 to 0.04 | <0.001 |
| PROMIS Anxiety | 0.67 | 0.66 to 0.68 | <0.001 | 0.67 | 0.66 to 0.68 | <0.001 |
| Age (per year) | 0.00 | -0.01 to 0.00 | 0.29 | 0.00 | -0.01 to 0.00 | 0.86 |
| Gender (Man) | -0.03 | -0.20 to 0.14 | 0.74 | -0.07 | -0.24 to 0.10 | 0.41 |
| Race (White) | 0.49 | 0.25 to 0.72 | <0.001 | 0.51 | 0.27 to 0.75 | <0.001 |
| Visit 2 | -0.16 | -2.48 to 2.16 | 0.89 | -0.16 | -1.74 to 1.42 | 0.84 |
| Visit 3 | -0.66 | -2.98 to 1.66 | 0.58 | -0.67 | -2.25 to 0.92 | 0.41 |
| **Random Effects** |  |  |  |  |  |  |
| $\sigma^2$ | 27.55 | - | - | 27.57 | - | - |
| $\tau_{00,id}$ | 10.82 | - | - | 10.88 | - | - |
| $\tau_{00,visit}$ | 0.70 | - | - | 0.32 | - | - |
| ICC | 0.29 | - | - | 0.29 | - | - |
| $N_{id}$ | 11,501 | - | - | 11,501 | - | - |
| $N_{visit}$ | 3 | - | - | 3 | - | - |
| Observations | 34,503 | - | - | 34,503 | - | - |
| Marginal $R^2$ / Conditional $R^2$ | 0.514 / 0.657 | - | - | 0.515 / 0.655 | - | - |

Abbreviations: PROMIS (Patient-Reported Outcomes Measurement Information System), NI (Not included in the model), $\sigma^2$ (random effect variance), $\tau_{00,id}$ (random intercept for each individual patient), $\tau_{00,visit}$ (random intercept for each clinic visit), ICC (Intra-Class-Correlation coefficient), $N_{id}$ (number of patients), $N_{visit}$ (number of clinic visits), Observations (number of datapoints for all included patients across all visits).

**Figure S1.**
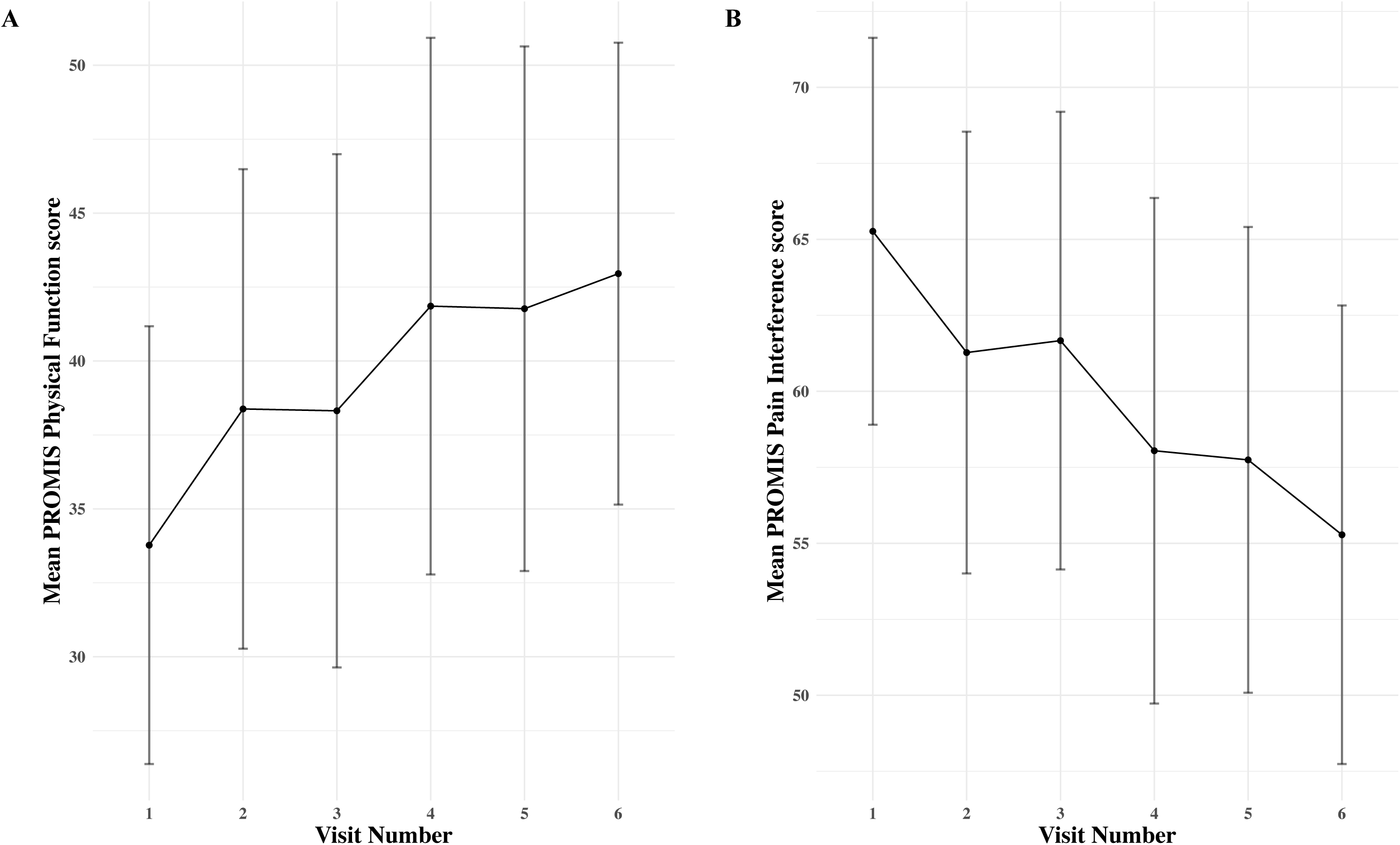
PROMIS physical health score changes among patients who achieved meaningful improvement. Mean PROMIS (A) Physical Function (N=1,672) and (B) Pain Interference (N=1,391) scores over time among patients who achieved meaningfully improved physical function or pain interference (of at least five PROMIS points) between their first and last clinic visit during the study period. Error bars represent one standard deviation. Abbreviation: PROMIS (Patient-Reported Outcomes Measurement Information System).

## References

1. Choi KW, Chen CY, Stein MB, et al. Assessment of Bidirectional Relationships Between Physical Activity and Depression Among Adults: A 2-Sample Mendelian Randomization Study. JAMA psychiatry. Apr 1 2019;76(4):399–408. doi:10.1001/jamapsychiatry.2018.4175

2. Hannerz H, Holtermann A, Madsen IEH. Musculoskeletal pain as a predictor for depression in the general working population of Denmark. Scand J Public Health. Aug 2021;49(6):589–597. doi:10.1177/1403494819875337

3. Guglielmo D, Hootman JM, Boring MA, et al. Symptoms of Anxiety and Depression Among Adults with Arthritis - United States, 2015-2017. MMWR Morb Mortal Wkly Rep. Oct 5 2018;67(39):1081–1087. doi:10.15585/mmwr.mm6739a2

4. Murphy LB, Sacks JJ, Brady TJ, Hootman JM, Chapman DP. Anxiety and depression among US adults with arthritis: prevalence and correlates. Arthritis Care Res (Hoboken). Jul 2012;64(7):968–76. doi:10.1002/acr.21685

5. Demyttenaere K, Bruffaerts R, Lee S, et al. Mental disorders among persons with chronic back or neck pain: results from the World Mental Health Surveys. Pain. Jun 2007;129(3):332–342. doi:10.1016/j.pain.2007.01.022

6. DeVeaugh-Geiss AM, West SL, Miller WC, Sleath B, Gaynes BN, Kroenke K. The adverse effects of comorbid pain on depression outcomes in primary care patients: results from the ARTIST trial. Pain medicine (Malden, Mass). 2010;11(5):732–741. doi:10.1111/j.1526-4637.2010.00830.x

7. Amtmann D, Cook KF, Jensen MP, et al. Development of a PROMIS item bank to measure pain interference. Pain. Jul 2010;150(1):173–82. doi:10.1016/j.pain.2010.04.025

8. Borges G, Aguilar-Gaxiola S, Andrade L, et al. Twelve-month mental health service use in six countries of the Americas: A regional report from the World Mental Health Surveys. Epidemiology and psychiatric sciences. Aug 27 2019;29:e53. doi:10.1017/s2045796019000477

9. Vranceanu AM, Beks RB, Guitton TG, Janssen SJ, Ring D. How do Orthopaedic Surgeons Address Psychological Aspects of Illness? The archives of bone andjoint surgery. 2017;5(1):2–9.

10. Weil TP. Insufficient dollars and qualified personnel to meet United States mental health needs. The Journal of nervous and mental disease. 2015;203(4):233–240. doi:10.1097/NMD.0000000000000271

11. Mojtabai R, Olfson M, Sampson NA, et al. Barriers to mental health treatment: results from the National Comorbidity Survey Replication. Psychol Med. Aug 2011;41(8):1751–61. doi:10.1017/s0033291710002291

12. Appelbaum PS, Parks J. Holding Insurers Accountable for Parity in Coverage of Mental Health Treatment. Psychiatr Serv. Feb 1 2020;71(2):202–204. doi:10.1176/appi.ps.201900513

13. Cheng AL, Leo AJ, Calfee RP, Dy CJ, Armbrecht MA, Abraham J. What Are Orthopaedic Patients’ and Clinical Team Members’ Perspectives Regarding Whether and How to Address Mental Health in the Orthopaedic Care Setting? A Qualitative Investigation of Patients With Neck or Back Pain. Clin Orthop Relat Res. Dec 8 2022;doi:10.1097/corr.0000000000002513

14. Beleckas CM, Guattery J, Chamberlain AM, Khan T, Kelly MP, Calfee RP. Using Patient-reported Outcomes Measurement Information System Measures to Understand the Relationship Between Improvement in Physical Function and Depressive Symptoms. J Am Acad Orthop Surg. Dec 15 2018;26(24):e511–e518. doi:10.5435/jaaos-d-17-00039

15. Zacharias AJ, Lemaster NG, Hawk GS, et al. Psychological Healthcare Burden Lessens After Hip Arthroscopy for Those With Comorbid Depression or Anxiety. Arthroscopy, sports medicine, and rehabilitation. Aug 2021;3(4):e1171–e1175. doi:10.1016/j.asmr.2021.05.005

16. Crijns TJ, Bernstein DN, Gonzalez R, Wilbur D, Ring D, Hammert WC. Operative Treatment is Not Associated with More Relief of Depression Symptoms than Nonoperative Treatment in Patients with Common Hand Illness. Clin Orthop Relat Res. Jun 2020;478(6):1319–1329. doi:10.1097/corr.0000000000001170

17. Reichman M, Bakhshaie J, Grunberg VA, Doorley JD, Vranceanu AM. What Are Orthopaedic Healthcare Professionals’ Attitudes Toward Addressing Patient Psychosocial Factors? A Mixed-Methods Investigation. Clin Orthop Relat Res. Feb 1 2022;480(2):248–262. doi:10.1097/corr.0000000000002043

18. Vranceanu AM, Bakhshaie J, Reichman M, Ring D. A Call for Interdisciplinary Collaboration to Promote Musculoskeletal Health: The Creation of the International Musculoskeletal Mental and Social Health Consortium (I-MESH). J Clin Psychol Med Settings. Oct 4 2021;doi:10.1007/s10880-021-09827-8

19. Ring D. Priorities for Advancing Mental and Social Health Among People Presenting for Care of Musculoskeletal Symptoms : International Consortium for Mental and Social Health in Musculoskeletal Care. J Clin Psychol Med Settings. Mar 22 2022;doi:10.1007/s10880-022-09865-w

20. Rose M, Bjorner JB, Becker J, Fries JF, Ware JE. Evaluation of a preliminary physical function item bank supported the expected advantages of the Patient-Reported Outcomes Measurement Information System (PROMIS). J Clin Epidemiol. 2008;61(1):17–33. doi:10.1016/j.jclinepi.2006.06.025

21. Hung M, Clegg DO, Greene T, Saltzman CL. Evaluation of the PROMIS physical function item bank in orthopaedic patients. JOrthopRes. Jun 2011;29(6):947–53. doi:10.1002/jor.21308

22. Crins MHP, Terwee CB, Ogreden O, et al. Differential item functioning of the PROMIS physical function, pain interference, and pain behavior item banks across patients with different musculoskeletal disorders and persons from the general population. Qual Life Res. Jan 2 2019;doi:10.1007/s11136-018-2087-x

23. Pilkonis PA, Choi SW, Reise SP, Stover AM, Riley WT, Cella D. Item banks for measuring emotional distress from the Patient-Reported Outcomes Measurement Information System (PROMIS(R)): depression, anxiety, and anger. Assessment. Sep 2011;18(3):263–83. doi:10.1177/1073191111411667

24. Schalet BD, Pilkonis PA, Yu L, et al. Clinical validity of PROMIS Depression, Anxiety, and Anger across diverse clinical samples. J Clin Epidemiol. Feb 27 2016;doi:10.1016/j.jclinepi.2015.08.036

25. Cella D, Yount S, Rothrock N, et al. The Patient-Reported Outcomes Measurement Information System (PROMIS): progress of an NIH Roadmap cooperative group during its first two years. Med Care. 2007;45(5 Suppl 1):S3–S11. doi:10.1097/01.mlr.0000258615.42478.55

26. Lee AC, Driban JB, Price LL, Harvey WF, Rodday AM, Wang C. Responsiveness and Minimally Important Differences for 4 Patient-Reported Outcomes Measurement Information System Short Forms: Physical Function, Pain Interference, Depression, and Anxiety in Knee Osteoarthritis. J Pain. Sep 2017;18(9):1096–1110. doi:10.1016/j.jpain.2017.05.001

27. Kroenke K, Yu Z, Wu J, Kean J, Monahan PO. Operating characteristics of PROMIS four-item depression and anxiety scales in primary care patients with chronic pain. Pain Med. Nov 2014;15(11):1892–901. doi:10.1111/pme.12537

28. Haug TT, Mykletun A, Dahl AA. The association between anxiety, depression, and somatic symptoms in a large population: the HUNT-II study. Psychosom Med. Nov-Dec 2004;66(6):845–851. doi:10.1097/01.psy.0000145823.85658.0c

29. Lee DJ, Calfee RP. The Minimal Clinically Important Difference for PROMIS Physical Function in Patients With Thumb Carpometacarpal Arthritis. Hand (New York, NY). Oct 18 2019:1558944719880025. doi:10.1177/1558944719880025

30. Sandvall B, Okoroafor UC, Gerull W, Guattery J, Calfee RP. Minimal Clinically Important Difference for PROMIS Physical Function in Patients With Distal Radius Fractures. J Hand Surg Am. Jun 2019;44(6):454–459.e1. doi:10.1016/j.jhsa.2019.02.015

31. Chen CX, Kroenke K, Stump TE, et al. Estimating minimally important differences for the PROMIS pain interference scales: results from 3 randomized clinical trials. Pain. Apr 2018;159(4):775–782. doi:10.1097/j.pain.0000000000001121

32. Amtmann D, Kim J, Chung H, Askew RL, Park R, Cook KF. Minimally important differences for Patient Reported Outcomes Measurement Information System pain interference for individuals with back pain. J Pain Res. 2016;9:251–5. doi:10.2147/jpr.s93391

33. Satterthwaite FE. An Approximate Distribution of Estimates of Variance Components. Biometrics Bulletin. 1946;2(6):110–114. doi:10.2307/3002019

34. Kuznetsova A, Brockhoff PB, Christensen RHB. lmerTest Package: Tests in Linear Mixed Effects Models. Journal of Statistical Software. 12/06 2017;82(13):1–26. doi:10.18637/jss.v082.i13

35. Benjamini Y, Hochberg Y. Controlling the False Discovery Rate: A Practical and Powerful Approach to Multiple Testing. Journal of the Royal Statistical Society: Series B (Methodological). 1995;57(1):289–300. doi:https://doi.org/10.1111/j.2517-6161.1995.tb02031.x

36. Cheng AL, Schwabe M, Doering MM, Colditz GA, Prather H. The Effect of Psychological Impairment on Outcomes in Patients With Prearthritic Hip Disorders: A Systematic Review and Meta-analysis. The American journal of sports medicine. 2019:363546519883246–363546519883246. doi:10.1177/0363546519883246

37. Browne JA, Sandberg BF, D’Apuzzo MR, Novicoff WM. Depression is associated with early postoperative outcomes following total joint arthroplasty: a nationwide database study. J Arthroplasty. Mar 2014;29(3):481–3. doi:10.1016/j.arth.2013.08.025

38. Rasouli MR, Menendez ME, Sayadipour A, Purtill JJ, Parvizi J. Direct Cost and Complications Associated With Total Joint Arthroplasty in Patients With Preoperative Anxiety and Depression. J Arthroplasty. Feb 2016;31(2):533–6. doi:10.1016/j.arth.2015.09.015

39. Harris AB, Marrache M, Puvanesarajah V, et al. Are preoperative depression and anxiety associated with patient-reported outcomes, health care payments, and opioid use after anterior discectomy and fusion? Spine J. Aug 2020;20(8): 1167–1175. doi:10.1016/j.spinee.2020.03.004

40. Forrest SJ, Siegert RJ, Krägeloh CU, Landon J, Medvedev ON. Generalizability theory distinguishes between state and trait anxiety. Psychol Assess. Nov 2021;33(11):1080–1088. doi:10.1037/pas0001060

